# A Predictive Model for the Evolution of COVID-19

**DOI:** 10.1101/2020.04.13.20063271

**Authors:** Rajneesh Bhardwaj

## Abstract

We predict the evolution of the COVID-19 pandemic in several countries by using a regression-based predictive model. In particular, the growth rate of the infection has been fitted as an exponential decay, as compared to a linear decay, reported previously. The model has been validated with the data of China and South Korea, where the pandemic is nearing to its end. The data of Italy, Germany, Spain, and Sweden show that the peak of the infection has reached i.e. a time when the new infections will start to decrease as compared to the previous day. The model predicts the approximate number of total infections at the end of the outbreak. The model prediction of the USA, and Brazil show that the peak will reach in the next two-three weeks. The total number of infections in the USA is estimated to be around 4 million by the model. The reported data of India show the start of disease evolution, with a large initial scatter in the growth rate. The possible peak date and the total number of infections are predicted using the data available.

## 1 Introduction

COVID-19 pandemic, caused by SARS-CoV-2 virus, has affected most of the countries around the world and has presented several challenges to humanity and science. A simple predictive model in this context can help to devise or modify policies by the governments while designing the mitigation measures and to judiciously use the healthcare infrastructure. While there are several predictive models reported previously (e.g. Refs. [1, 2, 3]), the present modeling effort is to develop a data-driven, extrapolation model. A previous model used a linear fit to the growth rate of the inflection, that could not predict the data accurately [1]. Here we build upon the model in Ref. [1] and use an exponential function to fit the growth rate of the infection.

## 2 Methods

The following ODE governs the evolution of the disease in a given human population [1]: 

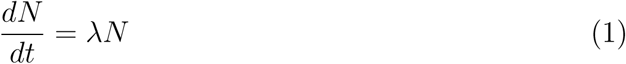

where *N* is number of people infected at a given time *t* and *λ* is the growth rate of the infections. The model uses reported field data of the infections of a population over a specific time period [*t*_0_, *t*_1_, *t*_2_, …….,*t*_*P*_], say [*N*_0_, *N*_1_, *N*_2_,…….,*N*_*p*_], where subscript 0 and *p* refers to the 0^*th*^ and present day of the outbreak. The growth rate can be numerically approximated from eq. 1 [1], 

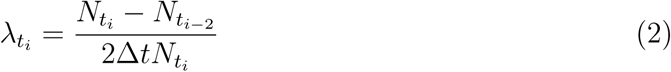

Considering the unit of time as day and the data is available for each day, we obtain Δ*t* = 1 in the above equation. We estimate 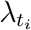 from the data of different countries given in public domain [4, 5, 6] and use a regression analysis based least squares fitting to the data of 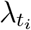. The fitting results in section 3 show that the exponential fit works well with the given data, as compared to a linear fit. Using the fit obtained, we extrapolate number of infections which will occur in future for *t* > *t*_*p*_ using the fitted value of 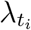. The initial value of this extrapolation is taken as *N*_*P*_. This allow us to predict the time of the peak of infections i.e. after this time, the daily infections will start to reduce. The total number of infections are also predicted, with the same extrapolation. The peak time is obtained by plotting 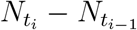 against time and the end of the outbreak is considered when 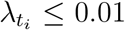 i.e. the rise in the total number of infections falls below 1%. The total infections are estimated at the end of outbreak.

## 3 Model validation

The model has been validated with the data of China and South Korea. These two counties have been selected since the outbreak of the epidemic is nearing to its end. Figs. 1 and 2 show that the time-history of the growth rate of the two countries and fitted exponential decays are in good agreement.

**Figure 1:**
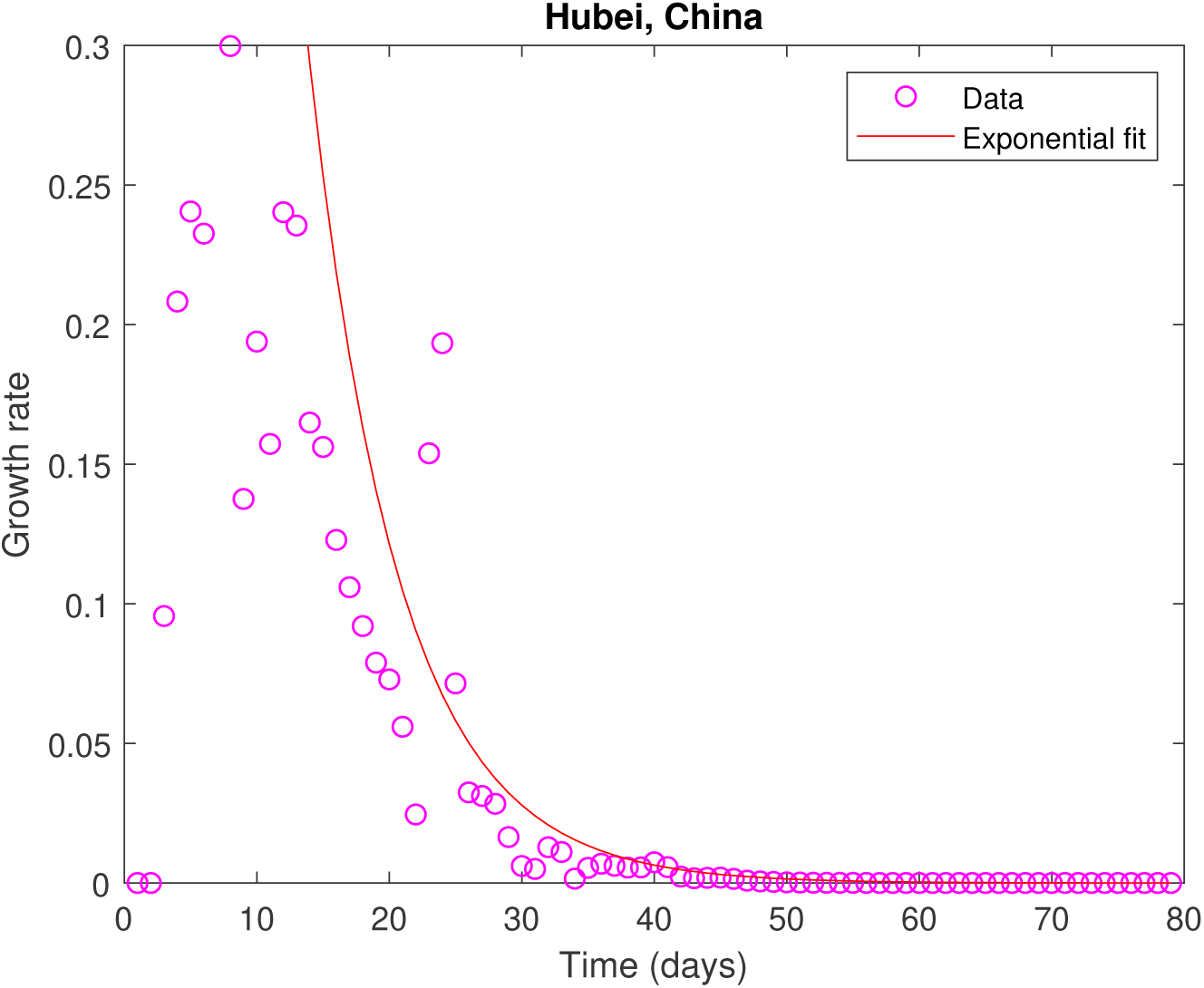
Validation of the model for with data of Hubei, China. Day 0 is 22 Jan, 2020. The growth rate is seen as an exponential decay, well-predicted by the model fit.

**Figure 2:**
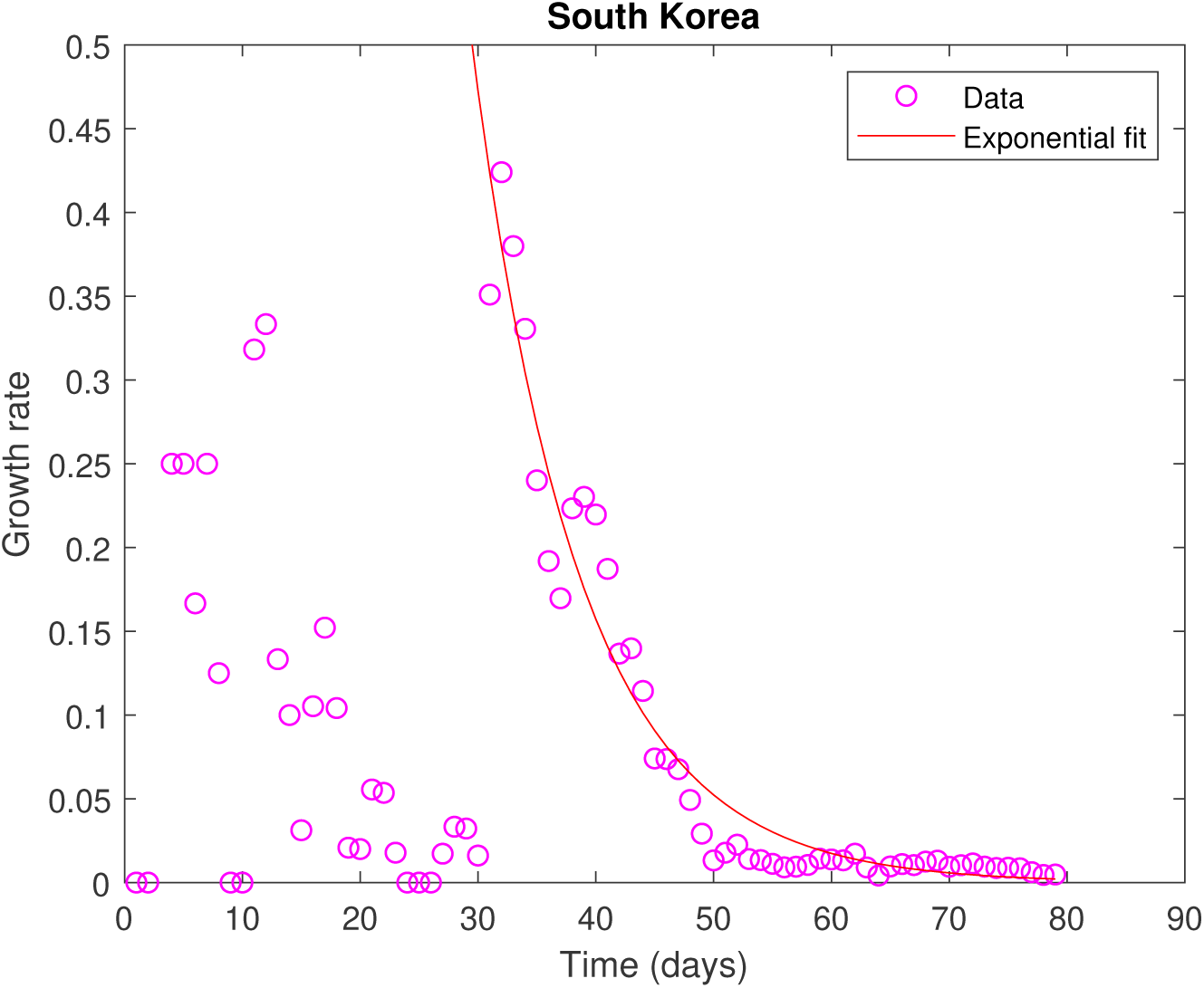
Validation of the model for South Korea. Rest of caption is the same as of Fig. 1.

## 4 Results and Discussion

First, we plot the results of the three countries - Italy (Fig. 3), Spain (Fig. 4), Germany (Fig. 5) and Sweden (Fig. 6), where the pandemic has reached its peak and the growth has slowed down. Time = 0 corresponds to 22 January 2020 in all figures. The peak date of the infection obtained from the data and the predicted number of infections have been listed for these countries in Table 1. The number of infections in Sweden will be one order of magnitude lesser than Italy, Germany, and Spain, despite no lock-down in the former.

**Table 1:** Predicted peak time of COVID-19 and total number of infections for different countries.

| Country | Date of peak | Total infections | Remarks |
| --- | --- | --- | --- |
| Italy | 24 Mar, 2020 | $2.8 \times 10^5$ | Lockdown |
| Germany | 3 Apr, 2020 | $2.5 \times 10^5$ | Partial lockdown |
| Spain | 2 Apr, 2020 | $2.6 \times 10^5$ | Partial lockdown |
| Sweden | 5 Apr, 2020 | $1.9 \times 10^4$ | No lockdown |
| USA | 03 May, 2020 | $4.2 \times 10^6$ | Partial lockdown |
| Brazil | 17 May, 2020 | $2.7 \times 10^5$ | Partial lockdown |
| India | 31 May, 2020 | $2.2 \times 10^5$ | Lockdown |

**Figure 3:**
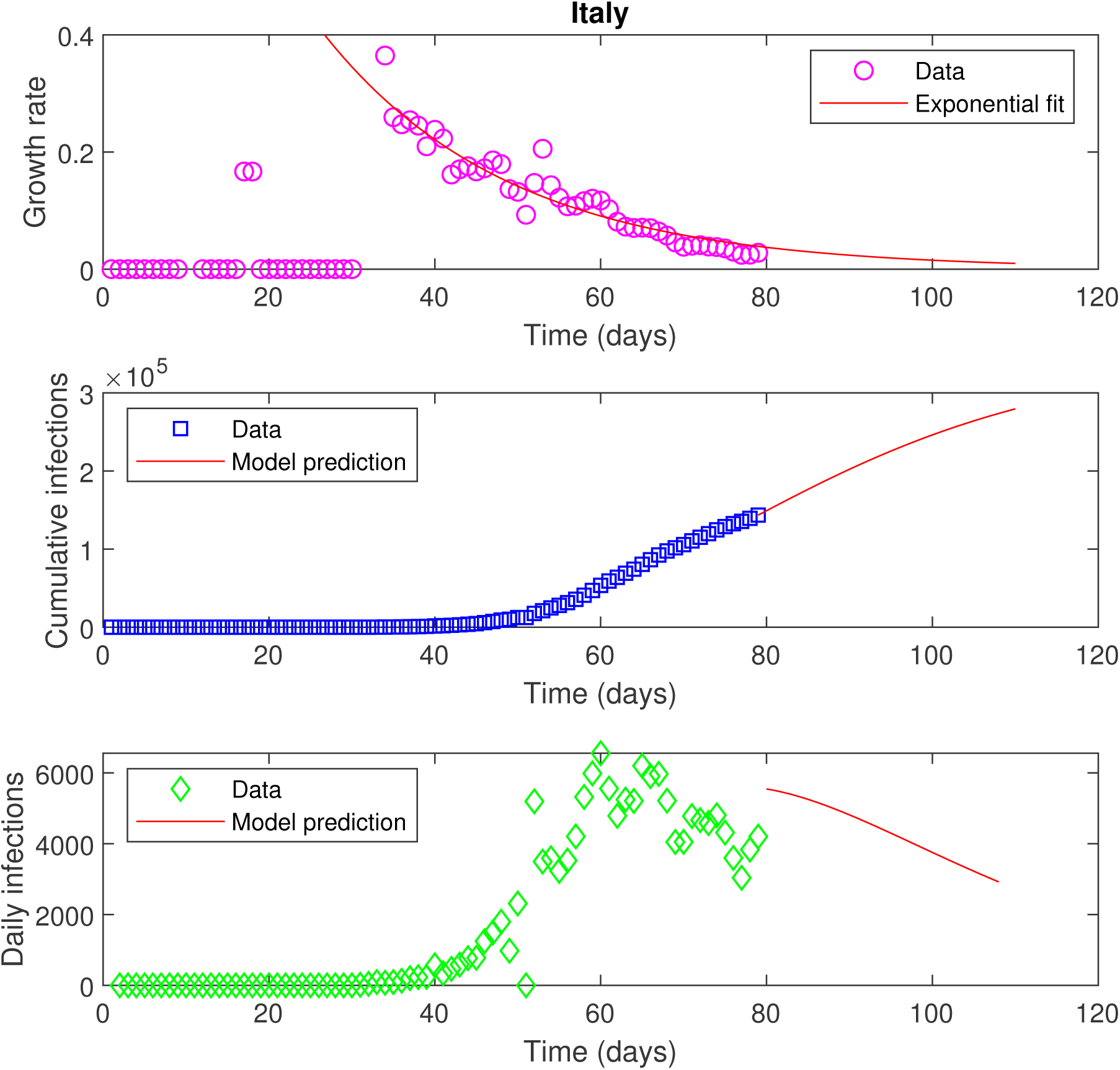
Data and model forecast for Italy. Day 0 is 22 Jan, 2020. Time-history of the growth rate of the infections per day (top row), cumulative number of infections (middle row) and daily infections (bottom row). The data is shown by symbols while the solid lines are predicted from the model. Simulation is stopped if 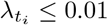 or the change in total infections per day is less than 1%.

**Figure 4:**
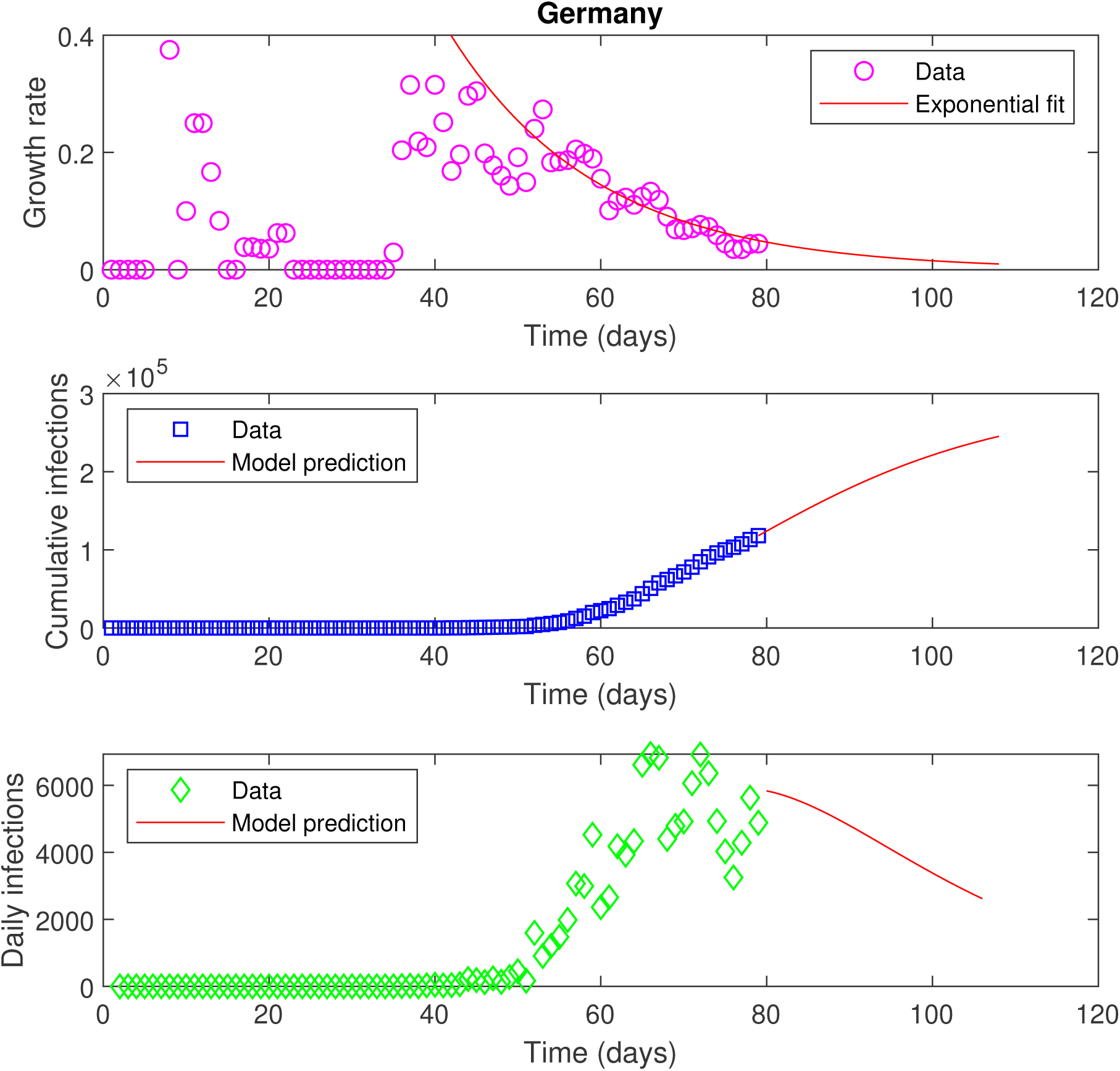
Data and model forecast for Germany. Rest of caption is the same as of Fig. 3.

**Figure 5:**
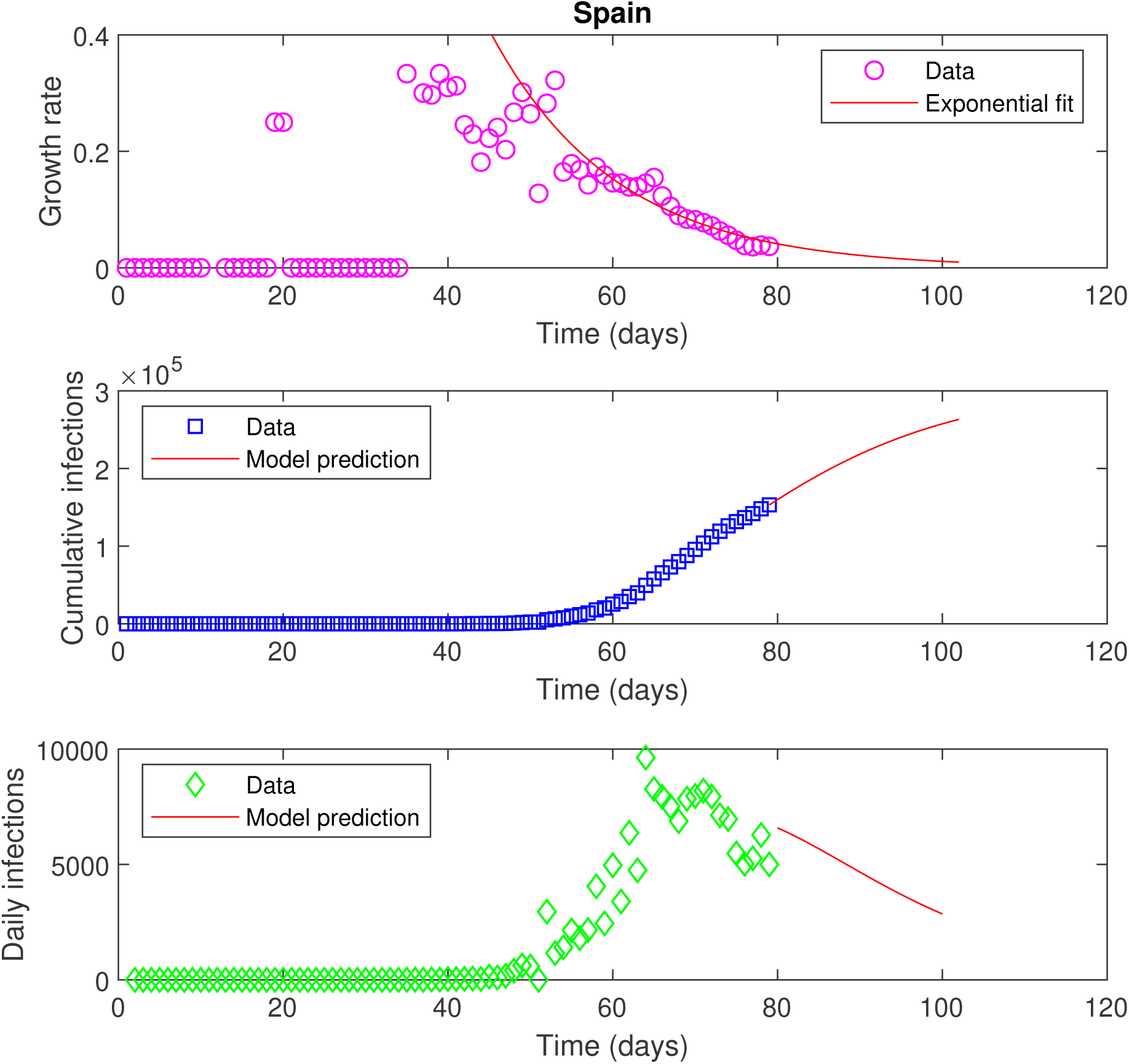
Data and model forecast for Spain. Rest of caption is the same as of Fig. 3.

**Figure 6:**
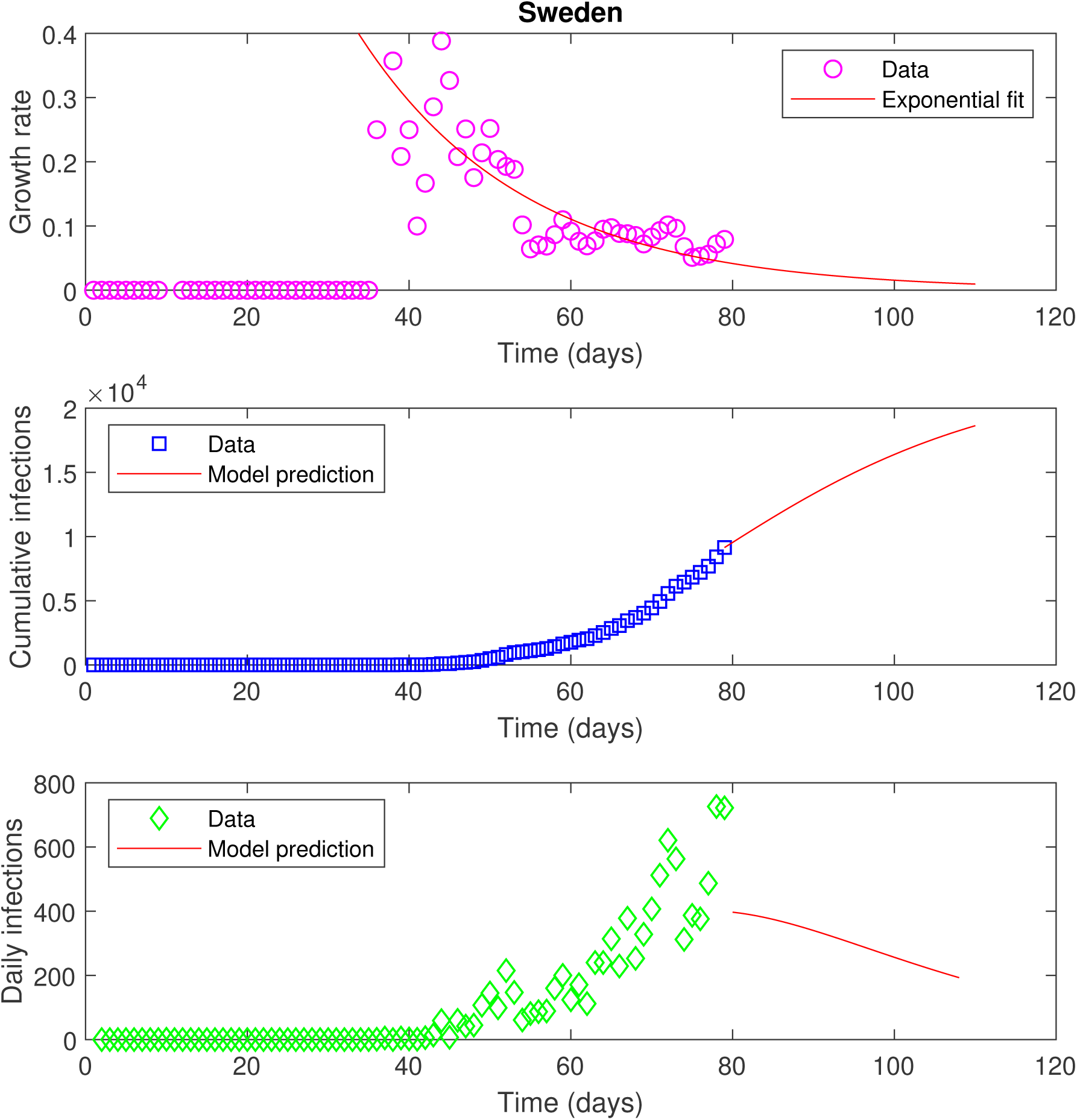
Data and model forecast for Sweden. Rest of caption is the same as of Fig. 3.

Second, we plot the results of countries that are expecting the peak of the infections in near future. Fig 7 plots the data of the USA and we can see that the peak of the epidemic is around 3 weeks away (from today, 11^*th*^ April 2020), with a total number of infections around 4.2*×*10^6^. The data of Brazil in Fig. 8 shows that the peak will reach after one week. The number of infections in Brazil will be one order of magnitude lesser than the USA.

**Figure 7:**
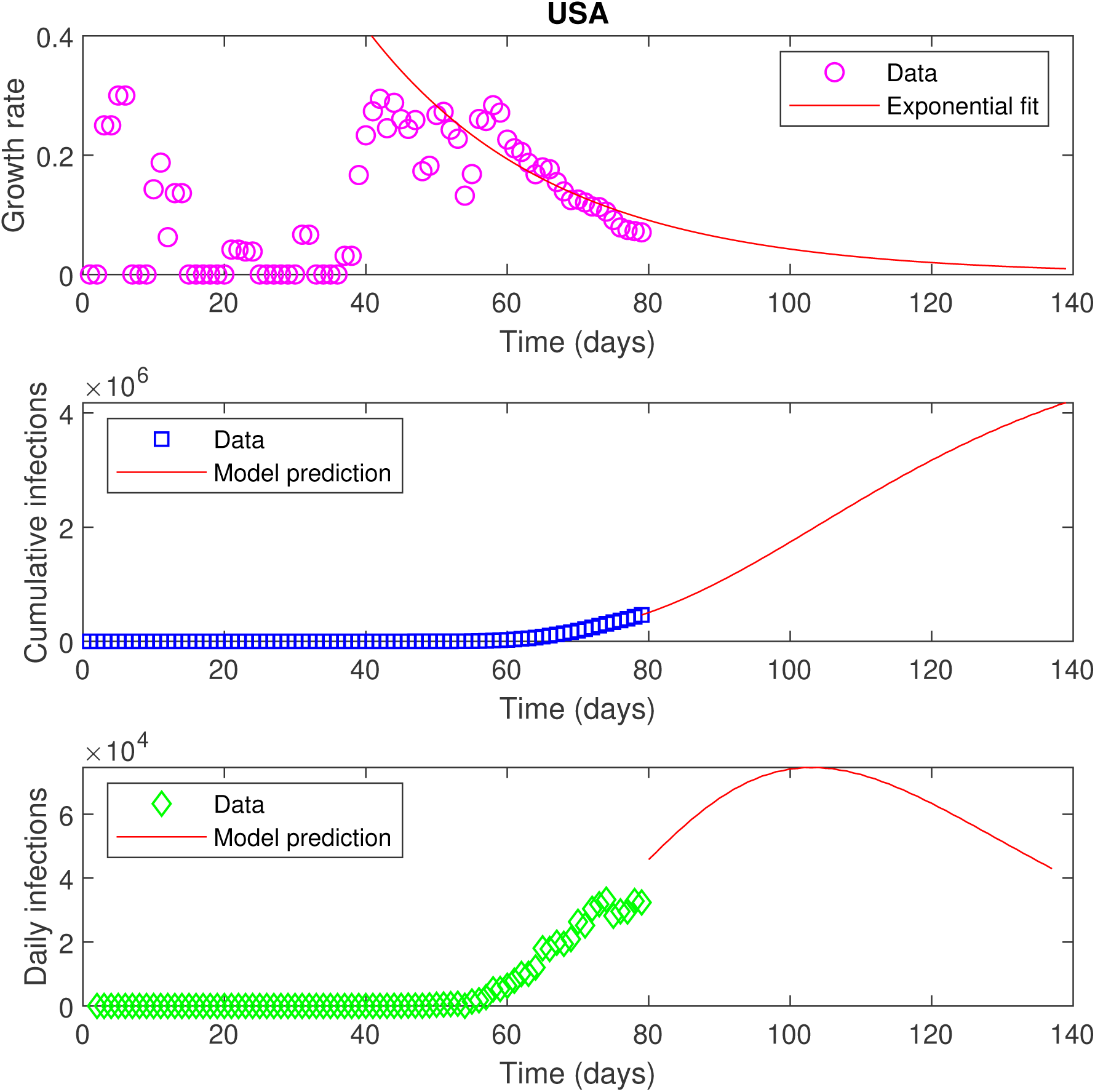
Data and model forecast for USA. Rest of caption is the same as of Fig. 3.

**Figure 8:**
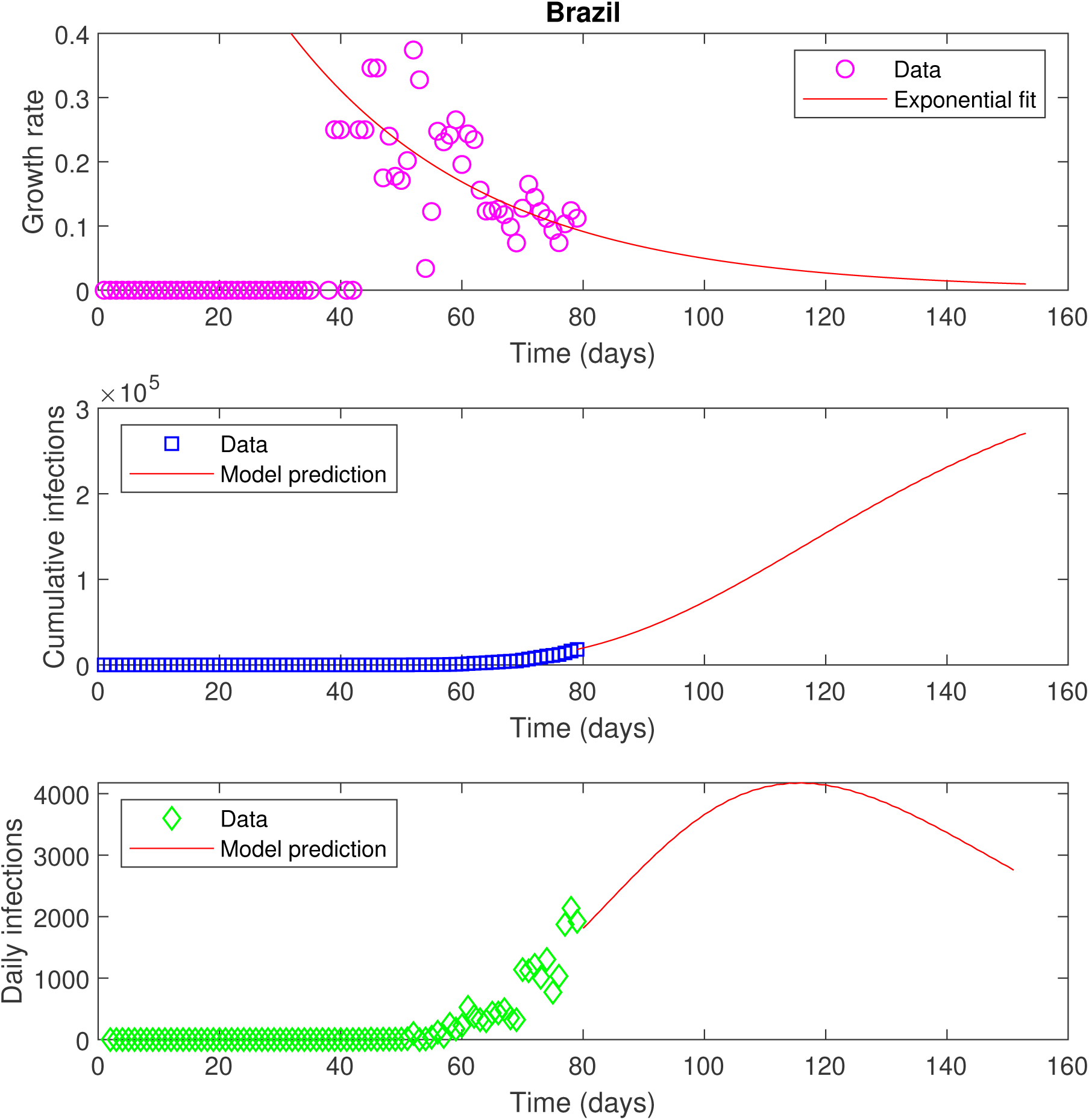
Data and model forecast for Brazil. Rest of caption is the same as of Fig. 3.

Finally, the data for India is plotted in Fig. 9. The data of growth rate in Fig 9 (first row) shows reasonable scatter and the *R*^2^ value of the exponential fit is low. The model prediction shows that the peak is around one and a half months away i.e on 31 May, 2020 (Table 1), with total infections on the order of 2.2 *×* 10^5^. A complete lock-down, imposed at the starting of the outbreak since 22 March 2020, maybe the reason for a slow outbreak. Since the model assumes uniform mixing, the predictions would become more accurate with time and the model forecast will improve with more data in the coming weeks.

**Figure 9:**
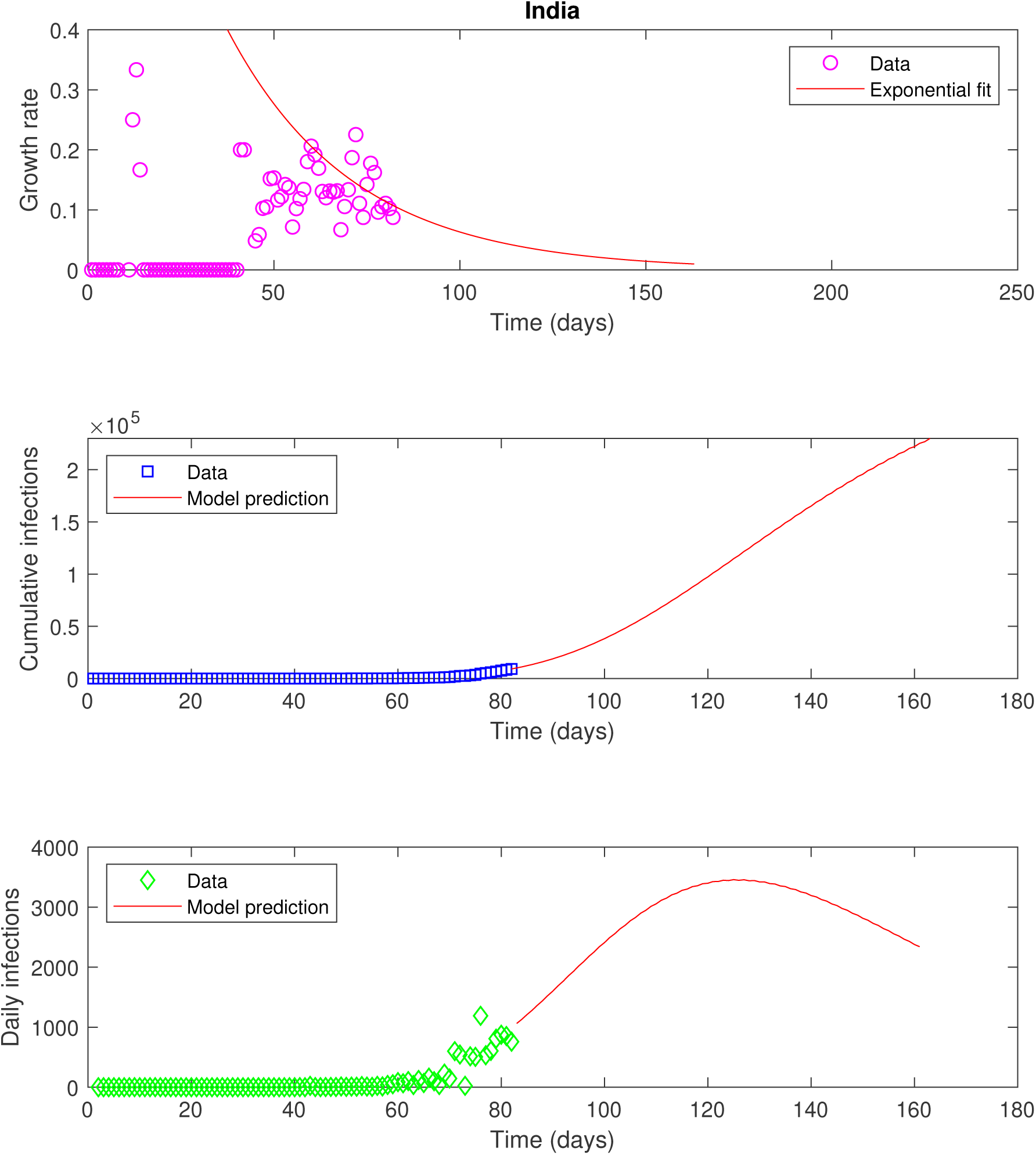
Data and model forecast for India. Rest of caption is the same as of Fig. 3.

Finally, the model presented here has certain limitations. It assumes a constant human population with uniform mixing of the people and cannot predict the total number of fatalities or recoveries. The model does not account for the mitigation measures, such as lock-down, taken by the respective governments. The model predictions are based on the reported data, that serves as an essential input to the model.

## Data Availability

The data used in the study is given in public domain on https://coronavirus.jhu.edu.

## 5 Acknowledgements

The work has been benefited from postings on online discussion forum of IITB faculty. I thank Prof. Amit Agrawal for useful discussions.

